# Distribution of cardiovascular disease risk based on the updated 2023 guideline-recommended Australian cardiovascular disease risk algorithm and comparison with the 2012 algorithm

**DOI:** 10.1101/2025.06.25.25330097

**Authors:** Nina Lazarevic, Meghana Bhat, Grace Joshy, Danielle C Butler, Mark Woodward, Anushka Patel, Rod Jackson, Garry Jennings, Rosemary Wyber, Ellie Paige, Emily Banks

## Abstract

**Objective:** Australian guidelines on cardiovascular disease (CVD) risk assessment and management, including risk prediction algorithms, were updated in 2023. We quantified CVD risk using the 2023 algorithm, compared this to the previous 2012 algorithm, and considered implications for preventive pharmacotherapy.

**Design:** Comparative analysis.

**Setting, participants:** Data from 115,873 people aged 45-74 years without existing CVD from MedicineInsight, a longitudinal primary-care database covering 8% of Australian general practices.

**Main outcomes:** We applied 2023 and 2012 risk algorithms to estimate and categorise individual-level CVD risk into low, intermediate, and high. Risk re-classification in accordance with the 2023 algorithm was not possible. Cohen’s kappa and Bland-Altman plots assessed agreement and concordance.

**Results:** Using the 2023 CVD risk algorithm and revised thresholds, 9.7% of participants were high (≥10% 5-year risk or clinically-determined high risk); 26.4% were intermediate (5-<10% 5-year risk), and 63.9% were low CVD risk (<5% 5-year risk). Corresponding 2012 figures were: 17.6% high (>15% 5-year risk or clinically-determined high risk); 11.6% intermediate (10-15% 5-year risk); and 70.8% low risk (<10% 5-year risk). Differences in proportions at high risk were largely driven by changes to clinically-determined high risk criteria. Overall, there was moderate-to-substantial agreement (kappa=0.62) and concordance (Kendall’s tau-b=0.74) between the algorithms.

**Conclusion:** Proportions estimated at low risk and not routinely recommended pharmacotherapy align with international standards and were similar between guidelines. Although fewer people would be recommended pharmacotherapy on the basis of high risk under the updated versus previous guidelines, this reflects better estimates of contemporary risk using the 2023 equation and does not account for re-classification. Pharmacotherapy is considered for those at intermediate risk depending on clinical context under the 2023 guidelines. To ensure continued reduction of the CVD burden across the population, we emphasise the application of re-classification factors and benefits of pharmacotherapy for those at intermediate risk.

**Summary box:** *The known:* Australian CVD prevention guidelines were updated in 2023, replacing 2012 guidelines. They recommend preventive pharmacotherapy for people at high 5-year CVD risk (≥10%) and considering it for intermediate risk (5-<10%).

*The new:* 9.7% of participants were classified as high, 26.4% as intermediate, and 63.9% as low CVD risk under the 2023 guidelines, using large-scale primary-care data. This compares with 17.6% high, 11.6% moderate and 70.8% low risk under 2012 guidelines.

*The implications:* Updated CVD risk algorithms more accurately reflect contemporary risk. Pharmacotherapy should be discussed systematically with people at intermediate CVD risk under the 2023 guidelines.

## Introduction

Cardiovascular disease (CVD), predominantly coronary heart disease and stroke, is a leading and highly preventable cause of death worldwide and in Australia.^1^ Interventions such as blood pressure-lowering and lipid-modifying therapies substantially lower the risk of CVD mortality and events and are World Health Organization Best Buys.^2-4^ In people without CVD, primary prevention practice guidelines in Australia and internationally recommend preventive treatment according to CVD risk, calculated using algorithms accounting for multiple risk factors and often including consideration of high-risk clinical conditions.^5,6^

In July 2023, new Australian primary CVD risk assessment and management guidelines were released, including a three-stage risk assessment algorithm, consisting of revised clinical high risk criteria, new CVD risk prediction equations, and risk reclassification criteria, and updated risk thresholds for initiating treatment.^5^ The new risk equations (referred to as AUS-PREDICT) are based on contemporary New Zealand PREDICT equations,^6,7^ re-calibrated for the Australian population.^8-10^ The 2023 guidelines replace the 2012 guidelines, which incorporated more extensive clinical high risk criteria and used the US Framingham risk equation,^11,12^ which is known to overestimate risk in the overall Australian population while underestimating risk in First Nations people.^8,9,13^ AUS-PREDICT includes additional risk predictors for area-level deprivation, history of atrial fibrillation, and CVD medication use. There is also an optional type-2 diabetes-specific equation (AUS-T2D-PREDICT) which includes additional diabetes-specific variables (Figure 1).^5^

**Figure 1.**
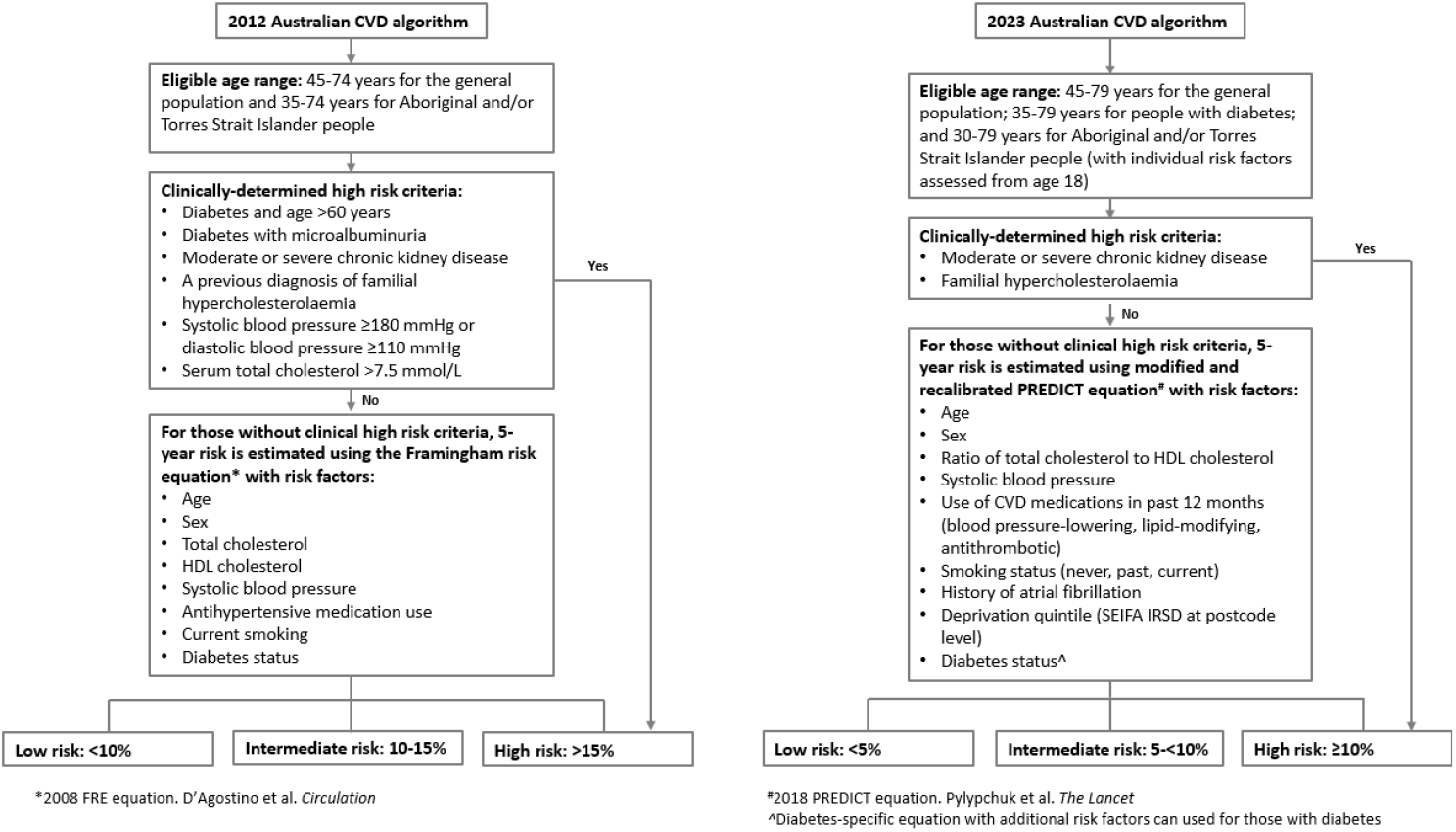
Eligibility, clinically-determined high risk criteria, and risk factors included in the 2023 and 2012 Australian CVD risk algorithms.

Under the 2023 Australian CVD risk guidelines, treatment with blood pressure- and lipid-modifying medication is recommended for those at high CVD risk—an estimated 5-year primary risk of ≥10%—with treatment considered for those at intermediate risk (5-<10%) depending on the clinical context. Risk categories and treatment thresholds were informed by a systematic review of international guidelines and modelling studies investigating preventive pharmacotherapy at different levels of CVD risk.^14^ Empirical data on the distribution of risk according to the updated 2023 guidelines and how they compare to the previous guidelines is important to inform implementation, including implications for ongoing primary CVD prevention in Australia.

The aim of this study was to quantify CVD risk distributions and classification into low, intermediate, and high risk under the 2023 guidelines, and to compare them with 2012 guidelines in a large Australian primary-care sample.

## Methods

### Data sources

We used the MedicineInsight database, established by NPS MedicineWise in 2011, containing longitudinal de-identified individual-level primary care data, extracted from general practice clinical information systems.^15^ The database contains patient information for over 3.2 million people, including sociodemographic data, prescribed medications, clinical measurements, and pathology results from 662 participating general practices across all states and territories, accounting for 8.2% of all Australian general practices.^15,16^ It is broadly representative of the Australian population based on age, sex, socioeconomic status, and the proportion of people identifying as Aboriginal and/or Torres Strait Islander.^16^

### Study population

The study population consisted of a 25% random sample of people in MedicineInsight who had at least one non-administrative clinical encounter provided by a doctor, nurse, or nurse practitioner between September 2020 and August 2022. To align with the recommended age range in the 2012 guidelines, we only included people aged 45–74 years. We excluded people who: had a prior history of documented CVD (including carotid artery stenosis, coronary heart disease, heart failure, peripheral vascular disease, renal artery stenosis, stroke, and transient ischaemic attack) or who had gestational diabetes.^17^

### Risk algorithms and outcomes

We conducted a head-to-head comparison of CVD risk estimated using the 2023 and 2012 risk algorithms, including the different thresholds for categorising risk (Figure 1).^5,11^

Outcomes were estimated CVD risk and the proportion of people in each CVD risk category (low, intermediate, and high). For risk assessment using the 2023 algorithm, we used AUS-T2D-PREDICT for people with type-2 diabetes who had additional diabetes-specific variables recorded and we used the general AUS-PREDICT otherwise. We also compared the use of AUS-PREDICT and AUS-T2D-PREDICT for those with type-2 diabetes to assess the impact of using the diabetes-specific equation. In a supplementary analysis, we used AUS-PREDICT for all people regardless of type-2 diabetes status.

### Risk factors

Risk factor definitions and approaches to dealing with missing data are detailed in Supplemental Table S1. We used risk factor measurements from the most recent clinical encounter within the study period (termed the “reference visit”), with allowable retrospective time windows for data measurements of one year for medications; two years for clinic measurements (blood pressure and body mass index (BMI)); three years for pathology data (glycated haemoglobin (HbA1c), urine albumin-creatinine ratio (uACR), estimated glomerular filtration rate (eGFR)), with the exception of total and high-density lipoprotein (HDL) cholesterol, for which we allowed five years. For pathology data, we also considered test results within one future month of the reference visit to be associated with the reference visit.

People with missing values for sex, age, smoking status, systolic blood pressure, or cholesterol were excluded. For the remaining risk factors, an absence of a specific disease diagnosis, prescription data, or pathology record was assumed to mean that the patient did not have that disease/medication. For the deprivation quintile,^18^ we imputed missing values with mean deprivation levels as per the 2023 guidelines.

### Statistical analysis

We first identified people who would be automatically considered at high risk based on existing clinical conditions. We then estimated primary CVD risk using the risk equations recommended within each guideline,^6,7,12^ and categorised participants as low, intermediate, or high risk.

After combining the high and clinically-determined high risk categories, we used the weighted kappa statistic to assess agreement between the risk classifications under the two algorithms, using linear weighting.

To assess agreement in risk estimates between the 2023 and 2012 risk equations, we used Bland-Altman plots of the differences versus means of the log-transformed risk estimates from the two equations.^19^ We used the Kendall’s tau-b statistic to quantify the discordance between the risk estimates. Analyses were performed in R v4.3.0.

### Ethics Approval

Ethics approval for this study was provided by the Australian National University Human Research Ethics Committee (reference 2021/424) and the Aboriginal Health & Medical Research Council of NSW (reference 1730/20).

## Results

### Sample characteristics

The study sample consisted of 115,873 people (Supplemental Figure S1). Just over half the study sample was female (54.5%, N=63,182), with an overall median age of 59 years (Q1: 52; Q3: 66 years), and 87.4% (N=101,227) were non- or past-smokers (Table 1). Few people had moderate-to-severe chronic kidney disease (1.1%), familial hypercholesterolemia (0.3%), or atrial fibrillation (2.6%). Within the twelve months before the reference visit, 35.8% of people had been prescribed blood pressure-lowering medications, 13.4% lipid-modifying medications, and 4.5% antithrombotic medications. Overall, 10.8% of people had type-2 diabetes (N=12,514), with a median 5 years since diagnosis (Q1: 2; Q3: 11 years), and 12.6% (1,575/12,514) of these were prescribed insulin-related medication (Supplemental Table S2).

**Table 1:** Patient characteristics.

|  | Female<br>(N=63,182) | Male<br>(N=52,691) | Overall<br>(N=115,873) |
| --- | --- | --- | --- |
| **Age (years)** |  |  |  |
| Median (Q1, Q3) | 60 (52,67) | 59 (52,66) | 59 (52,66) |
| **Age group (years)** |  |  |  |
| 45-54 | 20,366 (32.2%) | 17,566 (33.3%) | 37,932 (32.7%) |
| 55-64 | 22,688 (35.9%) | 19,140 (36.3%) | 41,828 (36.1%) |
| 65-74 | 20,128 (31.9%) | 15,985 (30.3%) | 36,113 (31.2%) |
| **Aboriginal and/or Torres Strait Islander person** |  |  |  |
| No | 61,972 (98.1%) | 51,779 (98.3%) | 113,751 (98.2%) |
| Yes | 1,210 (1.9%) | 912 (1.7%) | 2,122 (1.8%) |
| **Smoking status (2012 definition)** |  |  |  |
| Current smoker | 8,185 (13.0%) | 8,624 (16.4%) | 16,809 (14.5%) |
| Non-smoker | 54,997 (87.0%) | 44,067 (83.6%) | 99,064 (85.5%) |
| **Smoking status (2023 definition)** |  |  |  |
| Current smoking | 7,030 (11.1%) | 7,616 (14.5%) | 14,646 (12.6%) |
| Past smoking | 25,906 (41.0%) | 23,645 (44.9%) | 49,551 (42.8%) |
| Never smoked | 30,246 (47.9%) | 21,430 (40.7%) | 51,676 (44.6%) |
| **Chronic kidney disease, moderate to severe (2023 definition)** |  |  |  |
| No | 62,561 (99.0%) | 52,084 (98.8%) | 114,645 (98.9%) |
| Yes | 621 (1.0%) | 607 (1.2%) | 1,228 (1.1%) |
| **Chronic kidney disease, moderate to severe (2012 definition)** |  |  |  |
| No | 62,459 (98.9%) | 51,985 (98.7%) | 114,444 (98.8%) |
| Yes | 723 (1.1%) | 706 (1.3%) | 1,429 (1.2%) |
| **Familial hypercholesterolemia** |  |  |  |
| No | 62,950 (99.6%) | 52,562 (99.8%) | 115,512 (99.7%) |
| Yes | 232 (0.4%) | 129 (0.2%) | 361 (0.3%) |
| **Systolic blood pressure (mmHg)** |  |  |  |
| Median (Q1, Q3) | 130 (120,141) | 135 (126,144) | 133 (123,142) |
| **Diastolic blood pressure (mmHg)** |  |  |  |
| Median (Q1, Q3) | 80 (73,87) | 81 (75,88) | 80 (74,87) |
| Missing | 37 (0.1%) | 27 (0.1%) | 64 (0.1%) |
| **Total cholesterol** |  |  |  |
| Median (Q1, Q3) | 5.4 (4.7,6.1) | 5.1 (4.4,5.8) | 5.2 (4.6,5.9) |
| **Total to high density lipoprotein cholesterol ratio** |  |  |  |
| Median (Q1, Q3) | 3.4 (2.9,4.2) | 4.1 (3.3,4.9) | 3.7 (3.0,4.5) |
| **Use of blood-pressure lowering medication** |  |  |  |
| No | 42,151 (66.7%) | 32,220 (61.1%) | 74,371 (64.2%) |
| Yes | 21,031 (33.3%) | 20,471 (38.9%) | 41,502 (35.8%) |
| **Use of lipid-modifying medication** |  |  |  |
| No | 55,624 (88.0%) | 44,742 (84.9%) | 100,366 (86.6%) |
| Yes | 7,558 (12.0%) | 7,949 (15.1%) | 15,507 (13.4%) |
| **Use of antithrombotic medication** |  |  |  |
| No | 60,837 (96.3%) | 49,794 (94.5%) | 110,631 (95.5%) |
| Yes | 2,345 (3.7%) | 2,897 (5.5%) | 5,242 (4.5%) |
| **History of atrial fibrillation** |  |  |  |
| No | 61,995 (98.1%) | 50,871 (96.5%) | 112,866 (97.4%) |
| Yes | 1,187 (1.9%) | 1,820 (3.5%) | 3,007 (2.6%) |
| **Socioeconomic quintile (SEIFA—IRSD, 2016)<sup>18</sup>** |  |  |  |
| 1 (most disadvantaged) | 10,970 (17.4%) | 9,320 (17.7%) | 20,290 (17.5%) |
| 2 | 13,012 (20.6%) | 10,902 (20.7%) | 23,914 (20.6%) |
| 3 | 12,909 (20.4%) | 10,872 (20.6%) | 23,781 (20.5%) |
| 4 | 11,784 (18.7%) | 10,052 (19.1%) | 21,836 (18.8%) |
| 5 (least disadvantaged) | 14,272 (22.6%) | 11,307 (21.5%) | 25,579 (22.1%) |
| Missing | 235 (0.4%) | 238 (0.5%) | 473 (0.4%) |
| **Diabetes mellitus type 2** |  |  |  |
| No | 57,368 (90.8%) | 45,991 (87.3%) | 103,359 (89.2%) |
| Yes | 5,814 (9.2%) | 6,700 (12.7%) | 12,514 (10.8%) |

People with missing risk factor data on smoking status, systolic blood pressure, and cholesterol had similar sex and deprivation quintile distribution as complete cases, and a similar proportion of Aboriginal and/or Torres Strait Islander people. However, complete cases included more older people and people with type-2 diabetes (Supplemental Table S3).

### Comparison of CVD risk classification

#### Overall study sample

Prior to applying the risk equations, 1.4% (1,586/115,873) of people were classified as having clinically-determined high risk according to the 2023 algorithm compared to 12.3% (14,195/115,873) using the 2012 algorithm (Figure 2). Applying the relevant equations to remaining participants, 8.3% (9,614/115,873) were estimated to be high risk, 26.4% (30,637/115,873) intermediate, and 63.9% (74,036/115,873) low using the 2023 algorithm, and 5.3% (6,198/115,873) were high, 11.6% (13,415/115,873) intermediate, and 70.8% (82,065/115,873) low using the 2012 risk algorithm (Figure 2).

**Figure 2.**
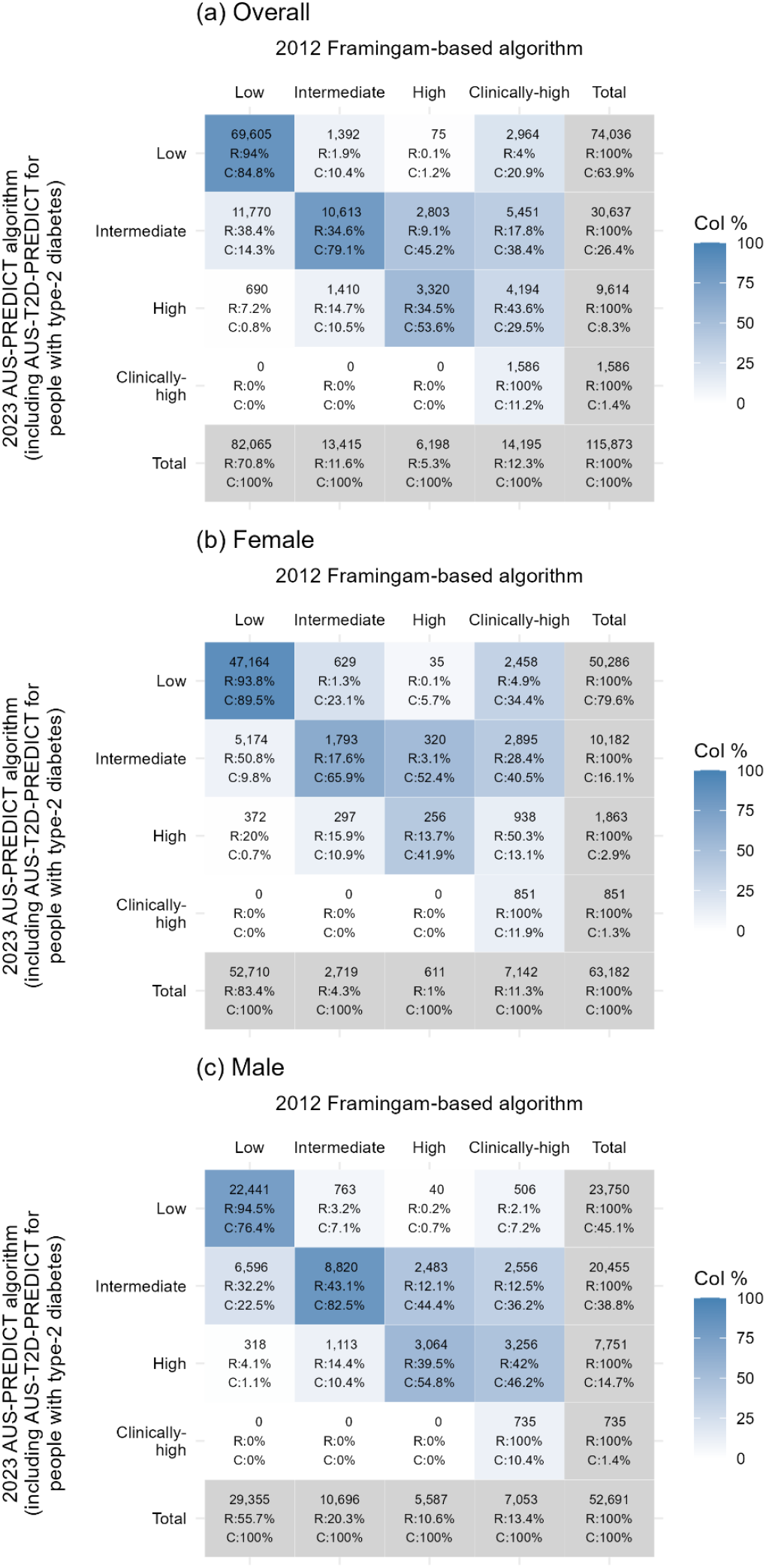
Risk classification under the 2023 and 2012 algorithms, overall and by sex, using AUS-T2D-PREDICT for people with type-2 diabetes and AUS-PREDICT otherwise. Row (R) and column (C) percentages are shown beneath cell frequencies; cell shading reflects the column percentage.

Combining clinically-determined and equation-derived risk, under the 2023 algorithm, 9.7% of participants were classified as high, 26.4% as intermediate, and 63.9% as low CVD risk. This compares with 17.6% high, 11.6% intermediate and 70.8% low risk under the 2012 algorithm (Figure 3). The pattern of greater proportions at intermediate risk with the updated algorithm was more pronounced in older age categories (Supplemental Figure S2).

**Figure 3.**
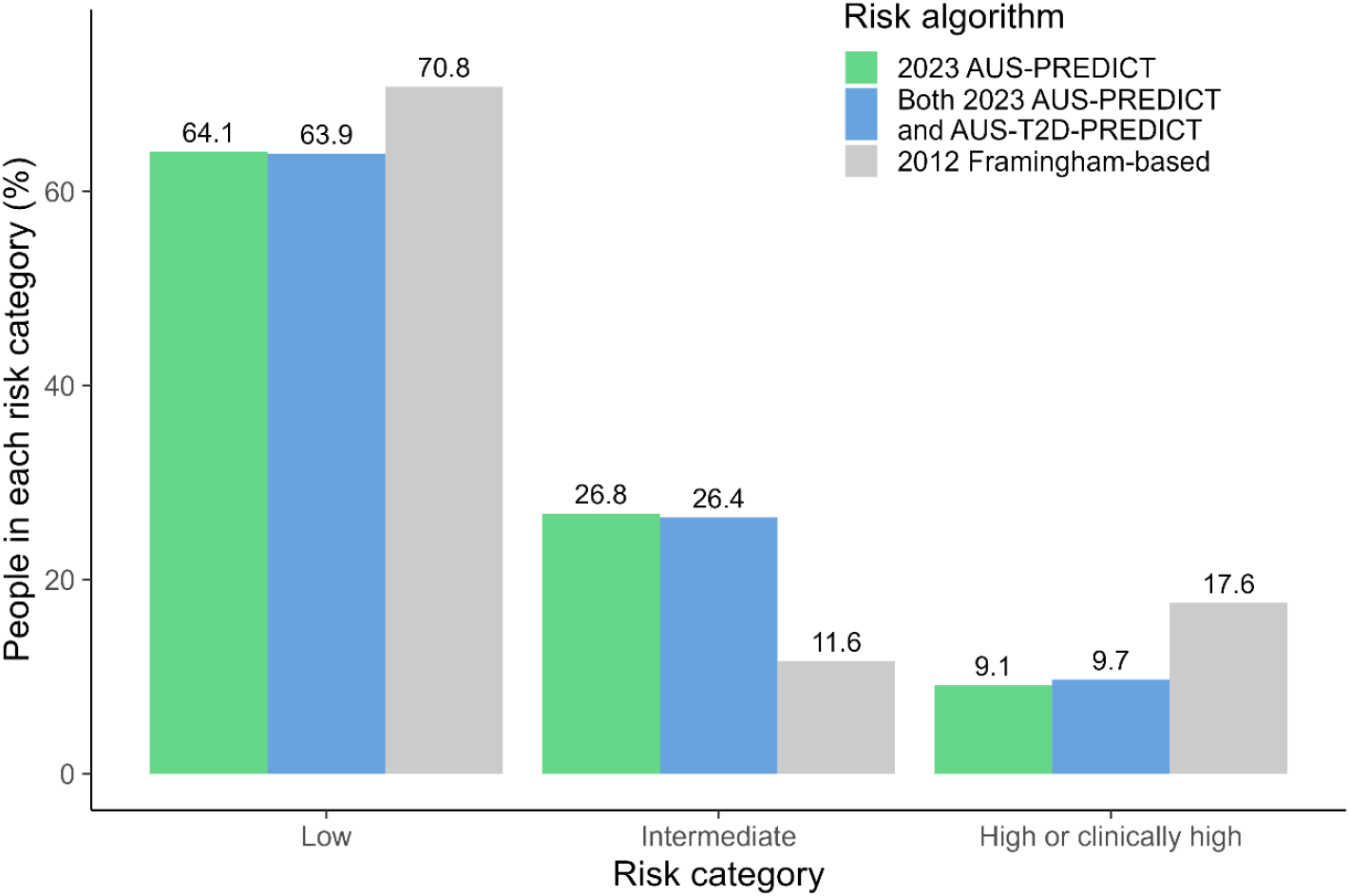
Risk classification under the 2023 and 2012 algorithms.

Of the people who were classified as low and high risk under the 2023 algorithm, 94.0% (69,605/74,036) and 81.3% (9,100/11,200) were also classified as low and high, respectively, under the 2012 algorithm. For people who were classified as of intermediate risk under the 2023 algorithm, 34.6% (10,613/30,637) were classified as of intermediate risk under the 2012 algorithm, while 38.4% (11,770/30,637) and 26.9% (8,254/30,637) were classified as of low and high risk, respectively, reflecting the pattern of movement to intermediate classification under the 2023 algorithm. (Figure 2). The proportions were similar when using the AUS-PREDICT general equation for all people, regardless of diabetes status (Supplemental Table S4).

Consistent with this, most people remained within the same risk category (77.1%; 89,318/115,873) or were classified in the adjacent lower (8.3%; 9,646/115,873) or upper category (11.4%; 13,180/115,873) under the 2023 algorithm compared to the 2012 algorithm (Figure 2), largely driven by the proportion at low risk being broadly similar between the two algorithms. Overall, 3.2% (3,729/115,873) of people moved up or down two categories. The proportion of agreement, after movement by chance was excluded, was 0.62 (weighted kappa; 95%CI 0.61 to 0.62), suggesting moderate-to-substantial agreement between the two algorithms for classifying risk.

When examining classification by sex, differences in classifications were more pronounced for females: of those classified at intermediate risk under the 2023 algorithm, 17.6% (1,793/10,182) were classified as intermediate and 50.8% (5,174/10,182) were classified as low risk under the 2012 algorithm (Figure 2 and Supplemental Table S4). Males were more likely to remain classified within the same risk category (e.g., of those classified as intermediate risk under the 2023 algorithm, 43.1% (8,820/20,455) were classified as intermediate risk under the 2012 algorithm).

#### Impact of the type-2 diabetes-specific equation under the 2023 guideline

Of the people diagnosed with type-2 diabetes, 61% (N=7,635/12,514) had additional diabetes-specific risk factor data recorded. When using the diabetes-specific rather than the general equation, most people with type-2 diabetes remained within the same risk category (72.1%; 5,502/7,635) or were classified in the adjacent lower (8.4%; 638/7,635) or upper category (19.1%; 1,459/7,635) (Supplemental Table S5). Notably, for those classified as high risk under AUS-T2D-PREDICT, 35.5% (977/2,755) would have been classified as intermediate under AUS-PREDICT.

### Comparison of CVD risk estimates using the 2023 and 2012 equations

Estimated median CVD risk was 3.7% (Q1: 2.0%, Q3: 6.2%) using the 2023 equation (AUS-T2D-PREDICT for people with type-2 diabetes and AUS-PREDICT otherwise) and was 5.3% (2.9%, 8.8%) using the 2012 Framingham-based equation. (Supplemental Figures S3). Values were generally lower for females than males (Supplemental Figures S4A and S4B).

The Bland-Altman plots suggested that the 2023 risk estimates were on average lower by 2.3% in absolute terms than the 2012 risk estimates (Supplemental Figure S5A). In relative terms, the 2023 risk estimates were on average 30% lower than the 2012 estimates (geometric mean) (Supplemental Figure S5B). Discrepancies between risk estimates from the two equations were smaller for very low and very high mean risk. The two equations were more likely to be concordant than discordant when ranking two people chosen at random in terms of their risk (Kendall’s tau-b statistic of 0.74).

## Discussion

In this large primary care population, almost 10% of people were estimated as high risk, where pharmacotherapy would be recommended. A further quarter of people were at intermediate risk and would be considered for treatment, depending on clinical context, under the latest guidelines. Around two-thirds would not routinely be recommended for preventive pharmacotherapy.

The 2023 equation was based on a contemporary New Zealand cohort of >400,000 people,^6,7^ recalibrated for Australia.^8-10^ The 2012 equation was derived from a 1970s Framingham Heart Study cohort of about 5,500 people,^20^ known to inaccurately predict risk for the Australian population. The 2023 equation more accurately reflects an individual’s contemporary risk of having a CVD event in the succeeding 5 years than the 2012 equation, with risks estimated for individuals being, on average, 30% lower than those calculated using the 2012 equation. However, this will have been counteracted, to some extent, by a lowering of the risk thresholds for categorising risk into low, intermediate, and high risk in the 2023 algorithm, based on best available evidence from systematic reviews and modelling, and aligning with major international guidelines.^14^

The 2023 algorithm included a narrower range of conditions leading to clinically-determined high risk than the 2012 risk algorithm, which rated individuals with common conditions such as diabetes with microalbuminuria as automatically at high CVD risk. This change is a more nuanced reflection of variation in CVD risk among people with specific conditions, particularly diabetes. Recent data advances allow risk to be quantified *within* these broad categories. Overall, the proportion of people automatically assigned as high risk based on clinical criteria in the updated guidelines was around an eighth of that observed under the 2012 algorithm. This has important implications for risk communication and shared decision making. For example, it means that people over 60 with well managed diabetes now have greater opportunity for individualised discussion of CVD risk and therapeutic options.

CVD assessment and management guidelines target preventive treatment to those at greater risk, while minimising overtreatment. There are currently large shortfalls in CVD preventive pharmacotherapy for people at high risk of future CVD.^21^ Applying the updated algorithm, more people would be categorised as being at intermediate risk than under the 2012 algorithm, and recommendations are that preventive medications be considered in this group, depending on clinical context. However, if only those at high risk are recommended treatment, 7.9% of people, who would have been recommended treatment under the 2012 risk guidelines, would not universally be recommended treatment under the 2023 guidelines (a 45% decrease).

The 2023 risk algorithm has a third stage which involves potential revising of risk categorisation for people with additional risk modifying factors, where the risk equations may less accurately predict risk. It is recommended clinicians consider upwards risk re-classification for: First Nations people, Māori people, Pacific Islander people, people of South Asian ethnicity, those with a family history of CVD, those without diabetes but with impaired renal function, and people with severe mental illness. Although we were unable to assess the impacts of risk re-classification, if, for example, half those at intermediate risk under the 2023 algorithm were re-classified to high risk based on the presence of risk-enhancing factors, then a larger proportion of people would be recommended preventive pharmacotherapy under the 2023 than the 2012 guidelines, resulting in greater reductions in CVD events across the population.

Clinical decision making about management of intermediate CVD risk should be carefully considered given the larger proportion of people now included in the intermediate category. In particular, the application of re-classification factors is likely to be important in further individualising risk assessments for people close to a category threshold. Blood pressure- and lipid-lowering medications lower the risk of future CVD events by 18-25% each, are effective at all levels of risk and have favourable safety profiles.^2,3^ Clinician education about implementation of the 2023 guidelines should continue to emphasise: 1) the application of re-classification factors; and, 2) the benefits of offering pharmacotherapy in the context of individualised decision making for people at intermediate risk.

This study is the first to our knowledge to quantify CVD risk and categorisation under the 2023 Australian CVD risk algorithm and to compare this to estimates using the previous 2012 algorithm. Strengths of this study include the use of a large-scale primary-care sample with general practice data from every state and territory in Australia; assessment and comparison of risk estimated using both clinically-determined high risk criteria and equations; and risk quantification using both the general equation and the diabetes-specific equation.

## Limitations

First, we restricted our analyses to people with recorded information on risk factors needed to calculate CVD risk as this best reflects what would occur in practice. Although our findings might be broadly indicative for the primary care population, caution should be used when considering how these results reflect CVD risk in the general Australian population. Comparisons of the proportions at risk between the algorithms are likely generalisable, but the absolute proportions are less likely to be generalisable. Second, analyses were restricted to people aged 45-74 years. The 2023 and 2012 guidelines differ in the age ranges eligible for assessment. Under the 2012 guidelines,^11^ people aged 45-74 years (30-74 for Aboriginal and Torres Strait Islander) were eligible for risk assessment.^22^ Under the 2023 guidelines, eligibility for risk assessment was extended to 79 years and people with diabetes are eligible from 35 years. Given that CVD risk increases substantially with increasing age, and that the age of risk assessment was extended in 2023, the proportions of people estimated to be at high risk using the 2023 algorithm will have been reduced by this age restriction. Finally, our study did not explicitly look at risk among First Nations people, who are at high risk of CVD earlier in life and have a greater CVD burden than non-Indigenous Australians.^23^ However, CVD risk algorithms used in Australian primary care to date have not been developed using First Nations data and have not been specifically calibrated for First Nations people. Comparison of the 2023 and 2012 CVD algorithms for First Nations people would be important to better understand changes in clinical practice and to address the inequitable burden of disease outcomes.

## Conclusions

The 2023 Australian CVD risk equation more accurately reflects contemporary risk than the previous equation. Applying the updated risk thresholds, pharmacotherapy would be recommended for around one tenth of this primary care population and considered for a further quarter, depending on clinical context. The major change is that more people will be calculated at intermediate risk than high risk. An unknown proportion of those people may be reassigned by clinicians to high risk based on re-classification factors. Therefore, the implementation and public health implications of the re-classification factors, particularly for people of intermediate risk, requires further consideration and research. Clinical decision making for people calculated at intermediate CVD risk is emerging as a key determinant of the impact of the 2023 guidelines update on future CVD events at the population level.

## Data Availability

The de-identified NPS MedicineWise data used for this study are not publicly available, but requests for the data can be made by contacting the Australian Commission on Safety and Quality in Health Care. The beta-coefficients for AUS-PREDICT and AUS-T2D-PREDICT used in this study for estimating CVD risk using the 2023 guidelines are not publicly available, but are available on application to the Heart Foundation of Australia.

## CRediT statement

Nina Lazarevic: Writing – original draft, Methodology, Formal analysis, Software, Visualization, Writing – review & editing; Meghana Bhat: Writing – original draft, Software, Visualization; Grace Joshy: Conceptualization, Methodology, Writing – review & editing; Danielle Butler: Writing – review & editing; Mark Woodward: Methodology, Writing – review & editing; Anushka Patel: Writing – review & editing; Rod Jackson: Writing – review & editing; Garry Jennings: Writing – review & editing; Rosemary Wyber: Writing – review & editing; Ellie Paige: Conceptualization, Writing – original draft, Visualization, Writing – review & editing. Emily Banks: Conceptualization, Validation, Writing – review & editing.

