## Supplemental for "Distribution of cardiovascular disease risk based on the updated 2023 guideline-recommended Australian cardiovascular disease risk algorithm and comparison with the 2012 algorithm"

### Table S1: Risk factor definitions and treatment of missing values.

| Risk factor | 2012 guideline | 2023 guideline |
|---|---|---|
| Deprivation quintile | N/A | We used the Australian Bureau of Statistics Index of Relative Socioeconomic Disadvantage (IRSD) 2016.[1] For two people who had a missing IRSD 2016 value but who had an IRSD 2011 value, we used the IRSD 2011 value. People with missing IRSD were set to the 2023 AUS-PREDICT equation mean deprivation levels, as per the AUS-PREDICT online calculator. |
| Diabetes status | We used an existing conditions field for diabetes status in the MedicineInsight database. The diagnostic algorithm was developed by NPS MedicineWise using diagnosis data and reasons recorded for the clinical encounter and prescriptions. The algorithm has been validated for type 2 diabetes,[2] with sensitivity of 0.89. We assumed that missing health condition data indicated absence or no history of the condition. | |
| Smoking status | Smoking status was defined using the latest clinician-recorded smoking status (current, ex-smoker, or non-smoker) and quit date where entered into the health record. For the 2012 risk equation, smoking status was defined as current smoker (including | Smoking status was defined using the latest clinician-recorded smoking status (current, ex-smoker, or non-smoker) and quit date where entered into the health record. For the 2023 risk equation, smoking status was defined as current smoker, past-smoker (including people |

<table>
<tr><td></td><td>people who stopped smoking within 12 months of the reference visit) or non-smoker (including people who stopped smoking >12 months prior to the reference visit).</td><td>who had a quit date before the reference visit, unless recorded as a current smoker at the reference visit), or never smoker (non-smokers with no recorded quit date).</td></tr>
<tr><td>Body mass index (BMI)</td><td>N/A</td><td>Weight (kg) divided by squared height (m), with weight limited to 35-250kg and height limited to 1-2.5m, and limited BMI to 15-60 kg/m$^2$. We used the most recently recorded BMI on or prior to the reference visit, with a maximum lookback of 2 years. BMI was used only in the AUS-T2D-PREDICT equation, which defaulted to AUS-PREDICT if any risk factors were missing.</td></tr>
<tr><td>Systolic and diastolic blood pressure</td><td colspan="2">We used the average of the last two seated measurements on two separate occasions, on and prior to the reference visit, with a maximum lookback period of 2 years. Observations of systolic blood pressure outside the range 30-300 mmHg, were set to missing. Individuals with missing systolic blood pressure data were not included in the analysis. For diastolic blood pressure, which is used only for determining 2012 clinical high risk, we assumed missing values meant lack of high diastolic blood pressure.</td></tr>
<tr><td>Total cholesterol, high-density lipoprotein (HDL) cholesterol, and the total:HDL cholesterol ratio</td><td colspan="2">We used the latest pathology test results within 1 month of the reference visit or up to 5 years prior, ignoring fasting status. HDL cholesterol values <0.25mmol/L and total cholesterol <0.5mmol/L were set to missing.[3,4] Individuals with missing lipids data were not included in the analysis.</td></tr>
<tr><td>Urine albumin-creatinine ratio (uACR), microalbuminuria, persistent proteinuria/macroalbuminuria</td><td colspan="2">For uACR, we used the latest pathology test results within 1 month of the reference visit or up to 3 years prior.<br>We defined microalbuminuria as uACR >2.5 mg/mmol for males and uACR >3.5 mg/mmol for females. We did not use the urinary albumin concentration to define microalbuminaria as it was not clear whether observations were spot samples or timed (24h) samples.<br>We defined persistent proteinuria/macroalbuminuria, as at least two positive test results within 3 months of the latest pathology test result associated with the reference visit. Positive results were defined as uACR >25 mg/mmol for males and uACR >35 mg/mmol for females.<br>Where uACR was missing, we assumed an absence of microalbuminuria. Where uACR was missing or was measured only once, we assumed an absence of persistent proteinuria/macroalbuminuria. For individuals with type 2 diabetes and missing uACR for use in the AUS-T2D-PREDICT equation, we used the AUS-PREDICT equation.</td></tr>
<tr><td>Estimated glomerular filtration rate (eGFR), low eGFR, sustained low eGFR</td><td colspan="2">We used the latest pathology test results within 1 month of the reference visit or up to 3 years prior. We defined low eGFR as eGFR <45 mL/min/1.73m$^2$, and sustained low eGFR as at least two observations with eGFR <45 mL/min/1.73m$^2$, using the latest two tests.<br>Although the AUS-PREDICT guidelines specify the use of the CKD-EPI formula, the MDRD formula was also allowed as it was used in a substantial number of cases. Where eGFR was recorded as a categorical rather than continuous value, we used the limit as the observed value (e.g., we used eGFR of 90 where ≥90 was recorded).<br>Where eGFR was missing, we assumed an absence of low eGFR. Where eGFR was missing or was only measured once, we assumed an absence of sustained low eGFR. For individuals with type 2 diabetes and missing eGFR for use in the AUS-T2D-PREDICT equation, we used the AUS-PREDICT equation.</td></tr>
<tr><td>Moderate-to-severe chronic kidney disease</td><td colspan="2">Moderate to severe chronic kidney disease was defined as either persistent proteinuria/macroalbuminuria (as defined above), or low eGFR (for the 2012 guidelines) or sustained low eGFR (for the 2023 guidelines, as defined above), or recorded stage 3-5 chronic kidney disease. Where none of these health states were recorded in the data, we assumed that this indicated an absence or no history of moderate to severe chronic kidney disease.</td></tr>
<tr><td>Glycated haemoglobin (HbA1c)</td><td>N/A</td><td>We used the latest pathology test results within 1 month of the reference visit or up to 3 years prior.<br>We set to missing HbA1c values outside the ranges 9-195 mmol/mol and 3-20% as per the online calculator at the National Glycohemoglobin Standardization Program website (https://ngsp.org/ifcc.asp).</td></tr>
</table>

| | | |
|---|---|---|
| | | HbA1c was used only in the AUS-T2D-PREDICT equation, which defaulted to AUS-PREDICT if any risk factors were missing. |
| Familial hypercholesterolemia | We identified cases of familial hypercholesterolemia by searching free text fields on diagnosis, clinical encounter, and prescription reasons for versions of familial hypercholesterolemia, familial dyslipidaemia, or familial hyperlipidaemia. Where we were not able to identify cases based on the recorded data, we assumed that this indicated absence or no history of the condition. | |
| Atrial fibrillation | N/A | We used an existing conditions field for atrial fibrillation in the MedicineInsight database. The diagnostic algorithm was developed by NPS MedicineWise using diagnosis data and reasons recorded for the clinical encounter and prescriptions.[2] We assumed that missing health condition data indicated absence or no history of the condition. |
| Lipid-modifying medications | Prescribed medications within one year of the reference visit, including those specified in the 2023 guidelines:<br>pravastatin, simvastatin, atorvastatin, fluvastatin, ezetimibe, acipimox, bezafibrate, cholestyramine, clofibrate, colestipol hydrochloride, gemfibrozil, and nicotinic acid. Absence of these prescriptions was considered to indicate absence of lipid-modifying medication use. | |
| Blood-pressure lowering medications | Prescribed medications within one year of the reference visit, including those specified in the 2023 guidelines:<br>angiotensin converting enzyme inhibitors (ATC codes C09A and C09B), betablockers (C07), thiazide (C03A), angiotensin II receptor blockers (C09C and C09D), and calcium channel blockers (C08). Absence of these prescriptions was considered to indicate absence of blood-pressure lowering medication use. | |
| Antithrombotic medications | Prescribed medications within one year of the reference visit, including those specified in the 2023 guidelines: aspirin (acetylsalicylic acid), clopidogrel, ticagrelor, dipyridamole, prasugrel, ticlopidine hydrochloride, warfarin, warfarin sodium, phenindione, dabigatran, and rivaroxaban. Absence of these prescriptions was considered to indicate absence of antithrombotic medication use. | |
| Insulin | N/A | Prescribed medications within one year of the reference visit, including insulins and analogues (ATC code A10A). Absence of these prescriptions was considered to indicate absence of insulin use. |
| Prior history of documented CVD (including carotid artery stenosis, coronary heart disease, heart failure, peripheral vascular disease, renal artery stenosis, stroke, and transient ischaemic attack) (exclusion criteria) | We used an existing conditions field in the MedicineInsight database. The diagnostic algorithm was developed by NPS MedicineWise using diagnosis data and reasons recorded for the clinical encounter and prescriptions.[2] We assumed that missing health condition data indicated absence or no history of the condition. | |
| Current gestational diabetes (exclusion criteria) | We used an existing conditions field in the MedicineInsight database. The diagnostic algorithm was developed by NPS MedicineWise using diagnosis data and reasons recorded for the clinical encounter and prescriptions.[2] We assumed that missing health condition data indicated absence of the condition. | |

### Figure S1: Flow chart outlining inclusions, exclusions, and final sample size

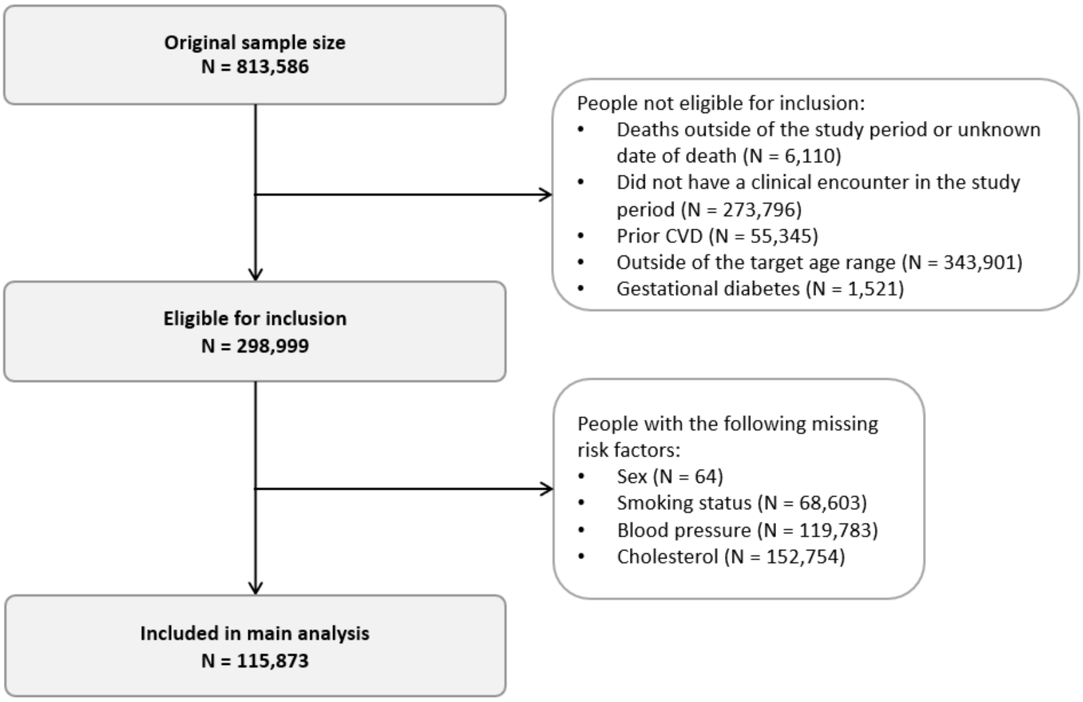


### Table S2: Summary statistics for diabetes-specific risk factors among people with type-2 diabetes.

| | Female (N=5,814) | Male (N=6,700) | Overall (N=12,514) |
|---|---|---|---|
| **Years since diagnosed with diabetes mellitus** | | | |
| Median (Q1, Q3) | 6.0 (2.0,11) | 5.0 (2.0,10) | 5.0 (2.0,11) |
| Missing N (%) | 106 (1.8%) | 111 (1.7%) | 217 (1.7%) |
| **Haemoglobin A1C (mmol/mol)** | | | |
| Median (Q1, Q3) | 51 (44,61) | 52 (45,62) | 52 (45,62) |
| Missing N (%) | 180 (3.1%) | 192 (2.9%) | 372 (3.0%) |
| **Urinary albumin to creatinine ratio** | | | |
| Median (Q1, Q3) | 1.2 (0.70,2.6) | 1.2 (0.70,3.4) | 1.2 (0.70,3.0) |
| Missing N (%) | 1,274 (21.9%) | 1,279 (19.1%) | 2,553 (20.4%) |
| **Estimated glomerular filtration rate (mL/min/1.73m$^2$)** | | | |
| Median (Q1, Q3) | 88 (74,90) | 89 (75,90) | 88 (75,90) |
| Missing N (%) | 216 (3.7%) | 282 (4.2%) | 498 (4.0%) |
| **Body mass index (kg/m$^2$)** | | | |
| Median (Q1, Q3) | 33 (28,38) | 31 (28,35) | 32 (28,36) |
| Missing N (%) | 1,090 (18.7%) | 1,286 (19.2%) | 2,376 (19.0%) |
| **Use of insulin-related medication** | | | |
| No | 5092 (87.6%) | 5847 (87.3%) | 10939 (87.4%) |
| Yes | 722 (12.4%) | 853 (12.7%) | 1575 (12.6%) |

Table S3: Distribution of selected risk factors among people who were not included in the analysis due to missing sex, smoking status, blood pressure, or cholesterol values.

| | Overall (N=183,126) |
|---|---|
| **Sex** | |
| Female | 100,551 (54.9%) |
| Male | 82,511 (45.1%) |
| Missing | 64 (0.0%) |
| **Age group** | |
| 45-54 | 75,756 (41.4%) |
| 55-64 | 62,236 (34.0%) |
| 65-74 | 45,134 (24.6%) |
| **Aboriginal and/or Torres Strait Islander person** | |
| No | 180,193 (98.4%) |
| Yes | 2,933 (1.6%) |
| **Socioeconomic quintile (SEIFA—IRSD, 2016)** | |
| 1 (most disadvantaged) | 29,963 (16.4%) |
| 2 | 38,000 (20.8%) |
| 3 | 36,641 (20.0%) |
| 4 | 36,623 (20.0%) |
| 5 (least disadvantaged) | 40,730 (22.2%) |
| Missing | 1169 (0.6%) |
| **Diabetes mellitus type 2** | |
| No | 177,057 (96.7%) |
| Yes | 6,069 (3.3%) |

Figure S2: Risk classification under the 2023 and 2012 algorithms, by sex and age category.

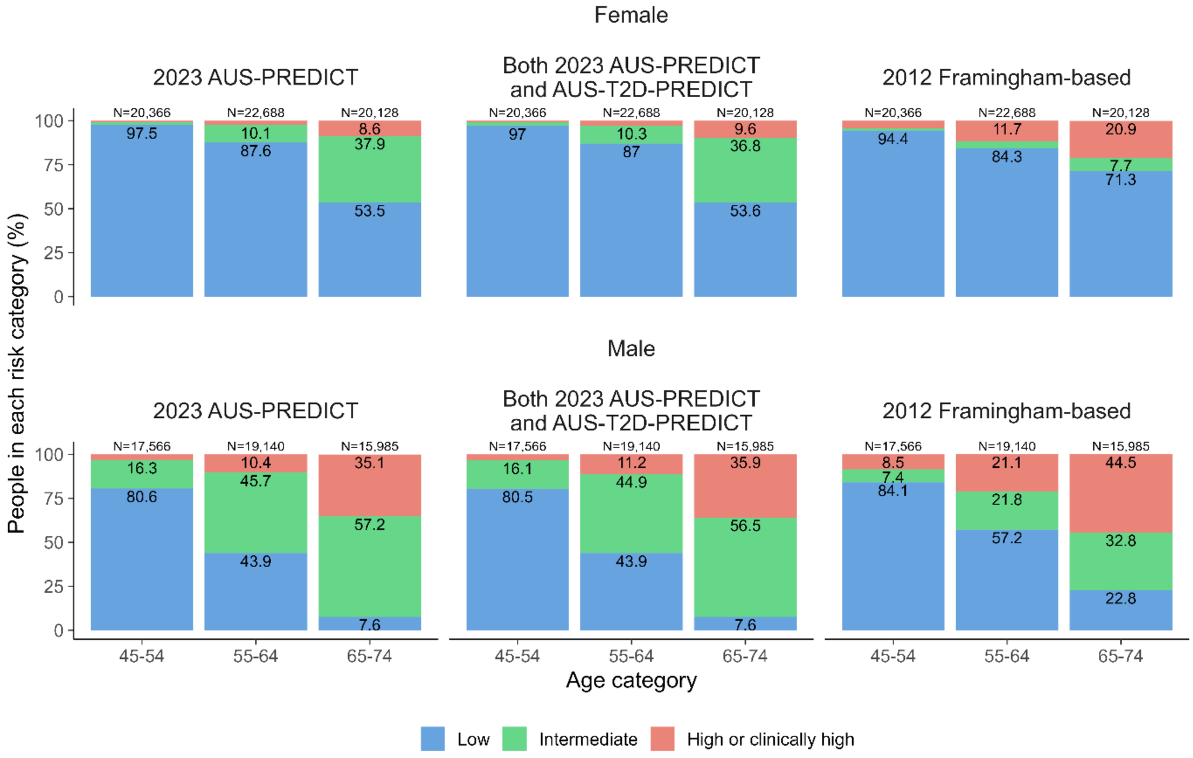

Table S4: Risk classification under the 2023 and 2012 algorithms, overall and by sex, using the AUS-PREDICT general equation regardless of diabetes status. Row (R) and column (C) percentages are shown beneath cell frequencies.

| 2023 AUS-PREDICT algorithm (general) | 2012 Framingham-based algorithm | | | | |
|---|---|---|---|---|---|
| OVERALL | **Low** | **Intermediate** | **High** | **Clinically High** | **Total** |
| **Low** | 69,731 | 1,396 | 74 | 3,085 | 74,286 |
| | *(R:93.9%) (C:85%)* | *(R:1.9%) (C:10.4%)* | *(R:0.1%) (C:1.2%)* | *(R:4.2%) (C:21.7%)* | *(R:100%) (C:64.1%)* |
| **Intermediate** | 11,674 | 10,641 | 2,758 | 5,957 | 31,030 |
| | *(R:37.6%) (C:14.2%)* | *(R:34.3%) (C:79.3%)* | *(R:8.9%) (C:44.5%)* | *(R:19.2%) (C:42%)* | *(R:100%) (C:26.8%)* |
| **High** | 660 | 1,378 | 3,366 | 3,567 | 8,971 |
| | *(R:7.4%) (C:0.8%)* | *(R:15.4%) (C:10.3%)* | *(R:37.5%) (C:54.3%)* | *(R:39.8%) (C:25.1%)* | *(R:100%) (C:7.7%)* |
| **Clinically High** | 0 | 0 | 0 | 1,586 | 1,586 |
| | *(R:0%) (C:0%)* | *(R:0%) (C:0%)* | *(R:0%) (C:0%)* | *(R:100%) (C:11.2%)* | *(R:100%) (C:1.4%)* |
| **Total** | 82,065 | 13,415 | 6,198 | 14,195 | 115,873 |
| | *(R:70.8%) (C:100%)* | *(R:11.6%) (C:100%)* | *(R:5.3%) (C:100%)* | *(R:12.3%) (C:100%)* | *(R:100%) (C:100%)* |
| FEMALE | **Low** | **Intermediate** | **High** | **Clinically High** | **Total** |
| **Low** | 47,281 | 653 | 35 | 2,539 | 50,508 |
| | *(R:93.6%) (C:89.7%)* | *(R:1.3%) (C:24%)* | *(R:0.1%) (C:5.7%)* | *(R:5%) (C:35.6%)* | *(R:100%) (C:79.9%)* |
| **Intermediate** | 5,065 | 1,788 | 335 | 3,089 | 10,277 |
| | *(R:49.3%) (C:9.6%)* | *(R:17.4%) (C:65.8%)* | *(R:3.3%) (C:54.8%)* | *(R:30.1%) (C:43.3%)* | *(R:100%) (C:16.3%)* |
| **High** | 364 | 278 | 241 | 663 | 1,546 |
| | *(R:23.5%) (C:0.7%)* | *(R:18%) (C:10.2%)* | *(R:15.6%) (C:39.4%)* | *(R:42.9%) (C:9.3%)* | *(R:100%) (C:2.4%)* |
| **Clinically High** | 0 | 0 | 0 | 851 | 851 |
| | *(R:0%) (C:0%)* | *(R:0%) (C:0%)* | *(R:0%) (C:0%)* | *(R:100%) (C:11.9%)* | *(R:100%) (C:1.3%)* |
| **Total** | 52,710 | 2,719 | 611 | 7,142 | 63,182 |
| | *(R:83.4%) (C:100%)* | *(R:4.3%) (C:100%)* | *(R:1%) (C:100%)* | *(R:11.3%) (C:100%)* | *(R:100%) (C:100%)* |
| MALE | **Low** | **Intermediate** | **High** | **Clinically High** | **Total** |
| **Low** | 22,450 | 743 | 39 | 546 | 23,778 |
| | *(R:94.4%) (C:76.5%)* | *(R:3.1%) (C:6.9%)* | *(R:0.2%) (C:0.7%)* | *(R:2.3%) (C:7.7%)* | *(R:100%) (C:45.1%)* |
| **Intermediate** | 6,609 | 8,853 | 2,423 | 2,868 | 20,753 |
| | *(R:31.8%) (C:22.5%)* | *(R:42.7%) (C:82.8%)* | *(R:11.7%) (C:43.4%)* | *(R:13.8%) (C:40.7%)* | *(R:100%) (C:39.4%)* |
| **High** | 296 | 1,100 | 3,125 | 2,904 | 7,425 |
| | *(R:4%) (C:1%)* | *(R:14.8%) (C:10.3%)* | *(R:42.1%) (C:55.9%)* | *(R:39.1%) (C:41.2%)* | *(R:100%) (C:14.1%)* |
| **Clinically High** | 0 | 0 | 0 | 735 | 735 |
| | *(R:0%) (C:0%)* | *(R:0%) (C:0%)* | *(R:0%) (C:0%)* | *(R:100%) (C:10.4%)* | *(R:100%) (C:1.4%)* |
| **Total** | 29,355 | 10,696 | 5,587 | 7,053 | 52,691 |
| | *(R:55.7%) (C:100%)* | *(R:20.3%) (C:100%)* | *(R:10.6%) (C:100%)* | *(R:13.4%) (C:100%)* | *(R:100%) (C:100%)* |

Table S5: Risk classification under the 2023 algorithm for people with type-2 diabetes only, overall and by sex, comparing the AUS-PREDICT general equation and the AUS-T2D-PREDICT equation that includes additional (diabetes-specific) risk factors. Row (R) and column (C) percentages are shown beneath cell frequencies.

| 2023 AUS-T2D-PREDICT algorithm | 2023 AUS-PREDICT algorithm | | | | |
|---|---|---|---|---|---|
| OVERALL | **Low** | **Intermediate** | **High** | **Clinically High** | **Total** |
| **Low** | 1,245<br>*(R:82.3%) (C:43.8%)* | 268<br>*(R:17.7%) (C:4.5%)* | 0<br>*(R:0%) (C:0%)* | 0<br>*(R:0%) (C:0%)* | 1,513<br>*(R:100%) (C:12.1%)* |
| **Intermediate** | 482<br>*(R:14.3%) (C:17%)* | 2,515<br>*(R:74.7%) (C:42.4%)* | 370<br>*(R:11%) (C:11.2%)* | 0<br>*(R:0%) (C:0%)* | 3,367<br>*(R:100%) (C:26.9%)* |
| **High** | 36<br>*(R:1.3%) (C:1.3%)* | 977<br>*(R:35.5%) (C:16.5%)* | 1,742<br>*(R:63.2%) (C:52.9%)* | 0<br>*(R:0%) (C:0%)* | 2,755<br>*(R:100%) (C:22%)* |
| **Clinically High** | 0<br>*(R:0%) (C:0%)* | 0<br>*(R:0%) (C:0%)* | 0<br>*(R:0%) (C:0%)* | 442<br>*(R:100%) (C:100%)* | 442<br>*(R:100%) (C:3.5%)* |
| **Missing T2D risk factor/s** | 1,078<br>*(R:24.3%) (C:37.9%)* | 2,176<br>*(R:49%) (C:36.7%)* | 1,183<br>*(R:26.7%) (C:35.9%)* | 0<br>*(R:0%) (C:0%)* | 4,437<br>*(R:100%) (C:35.5%)* |
| **Total** | 2,841<br>*(R:22.7%) (C:100%)* | 5,936<br>*(R:47.4%) (C:100%)* | 3,295<br>*(R:26.3%) (C:100%)* | 442<br>*(R:3.5%) (C:100%)* | 12,514<br>*(R:100%) (C:100%)* |
| FEMALE | **Low** | **Intermediate** | **High** | **Clinically High** | **Total** |
| **Low** | 1,059<br>*(R:85.7%) (C:45%)* | 176<br>*(R:14.3%) (C:6.6%)* | 0<br>*(R:0%) (C:0%)* | 0<br>*(R:0%) (C:0%)* | 1,235<br>*(R:100%) (C:21.2%)* |
| **Intermediate** | 369<br>*(R:23.5%) (C:15.7%)* | 1,128<br>*(R:72%) (C:42.4%)* | 70<br>*(R:4.5%) (C:11.8%)* | 0<br>*(R:0%) (C:0%)* | 1,567<br>*(R:100%) (C:27%)* |
| **High** | 29<br>*(R:4.1%) (C:1.2%)* | 358<br>*(R:51.1%) (C:13.5%)* | 313<br>*(R:44.7%) (C:52.8%)* | 0<br>*(R:0%) (C:0%)* | 700<br>*(R:100%) (C:12%)* |
| **Clinically High** | 0<br>*(R:0%) (C:0%)* | 0<br>*(R:0%) (C:0%)* | 0<br>*(R:0%) (C:0%)* | 212<br>*(R:100%) (C:100%)* | 212<br>*(R:100%) (C:3.6%)* |
| **Missing T2D risk factor/s** | 894<br>*(R:42.6%) (C:38%)* | 996<br>*(R:47.4%) (C:37.5%)* | 210<br>*(R:10%) (C:35.4%)* | 0<br>*(R:0%) (C:0%)* | 2,100<br>*(R:100%) (C:36.1%)* |
| **Total** | 2,351<br>*(R:40.4%) (C:100%)* | 2,658<br>*(R:45.7%) (C:100%)* | 593<br>*(R:10.2%) (C:100%)* | 212<br>*(R:3.6%) (C:100%)* | 5,814<br>*(R:100%) (C:100%)* |
| MALE | **Low** | **Intermediate** | **High** | **Clinically High** | **Total** |
| **Low** | 186<br>*(R:66.9%) (C:38%)* | 92<br>*(R:33.1%) (C:2.8%)* | 0<br>*(R:0%) (C:0%)* | 0<br>*(R:0%) (C:0%)* | 278<br>*(R:100%) (C:4.1%)* |
| **Intermediate** | 113<br>*(R:6.3%) (C:23.1%)* | 1,387<br>*(R:77.1%) (C:42.3%)* | 300<br>*(R:16.7%) (C:11.1%)* | 0<br>*(R:0%) (C:0%)* | 1,800<br>*(R:100%) (C:26.9%)* |
| **High** | 7<br>*(R:0.3%) (C:1.4%)* | 619<br>*(R:30.1%) (C:18.9%)* | 1,429<br>*(R:69.5%) (C:52.9%)* | 0<br>*(R:0%) (C:0%)* | 2,055<br>*(R:100%) (C:30.7%)* |
| **Clinically High** | 0<br>*(R:0%) (C:0%)* | 0<br>*(R:0%) (C:0%)* | 0<br>*(R:0%) (C:0%)* | 230<br>*(R:100%) (C:100%)* | 230<br>*(R:100%) (C:3.4%)* |
| **Missing T2D risk factor/s** | 184<br>*(R:7.9%) (C:37.6%)* | 1,180<br>*(R:50.5%) (C:36%)* | 973<br>*(R:41.6%) (C:36%)* | 0<br>*(R:0%) (C:0%)* | 2,337<br>*(R:100%) (C:34.9%)* |
| **Total** | 490<br>*(R:7.3%) (C:100%)* | 3,278<br>*(R:48.9%) (C:100%)* | 2,702<br>*(R:40.3%) (C:100%)* | 230<br>*(R:3.4%) (C:100%)* | 6,700<br>*(R:100%) (C:100%)* |

Figure S3: Distributions of CVD risk estimates under the 2023 and 2012 algorithms. Noting that the two 2023 risk estimates distributions (in blue and green) lie one top of each other.

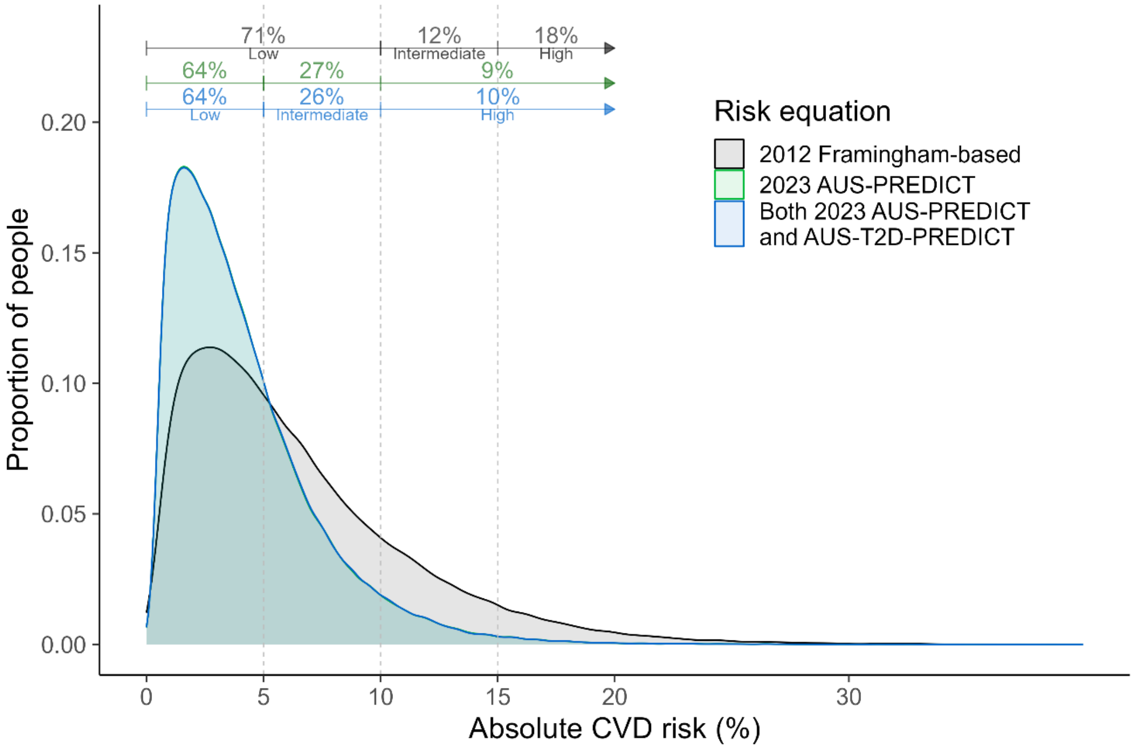


Note: The proportions in each risk category include people who were at clinically-determined high risk.

Figure S4A and S4B: Distributions of CVD risk estimates under the 2023 and 2012 algorithms, for females (A) and males (B). Noting that the two 2023 risk estimates distributions (in blue and green) lie one top of each other.

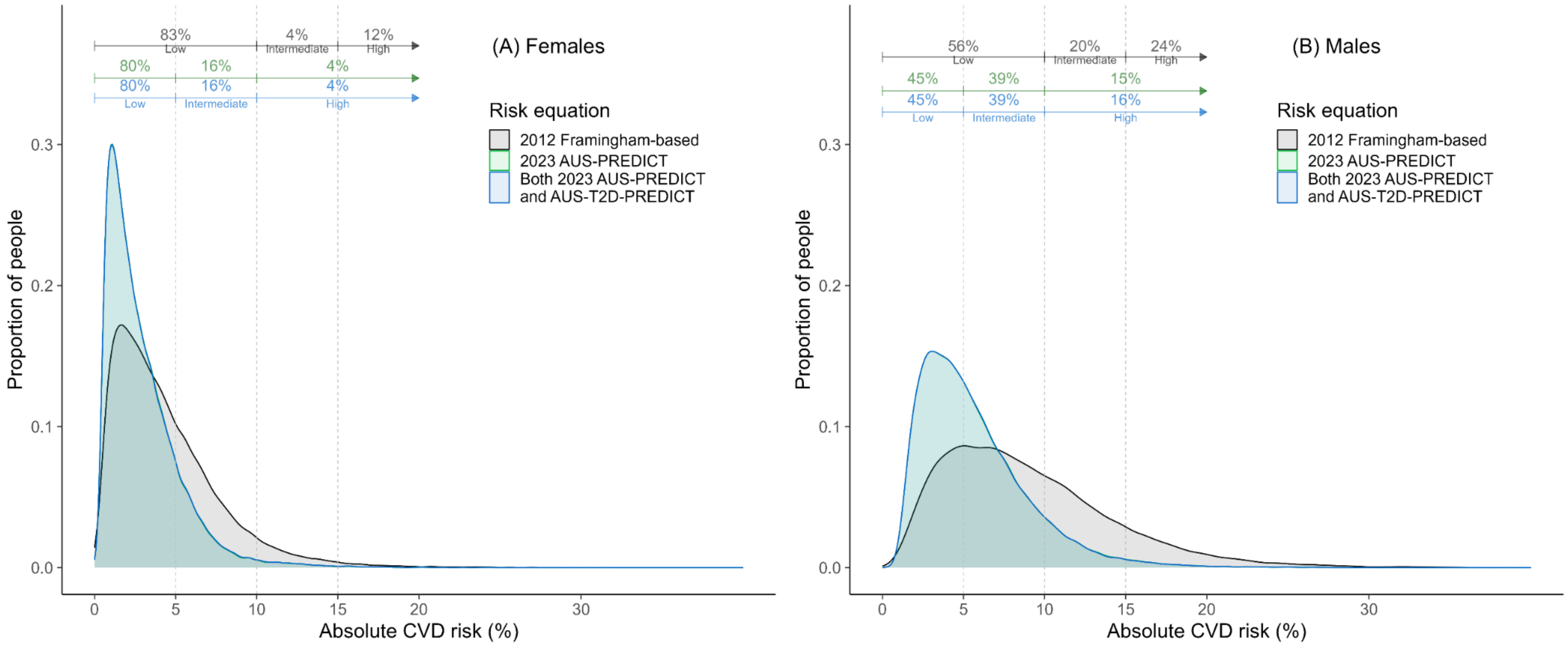


Note: The proportions in each risk category include people who were at clinically-determined high risk.

Figure S5A and S5B: Bland-Altman plots of 2023 AUS-PREDICT (including AUS-T2D-PREDICT for patients with type-2 diabetes) versus 2012 Framingham-based CVD risk estimates. Panel (A) shows the differences in risks and Panel (B) shows the difference in log risks (i.e., ratio of risks), versus the mean CVD risk across the two equations. Both plots show the bias (discrepancy). Panel B shows the lower and upper limits of agreement (LOA), within which 95% of differences lie.

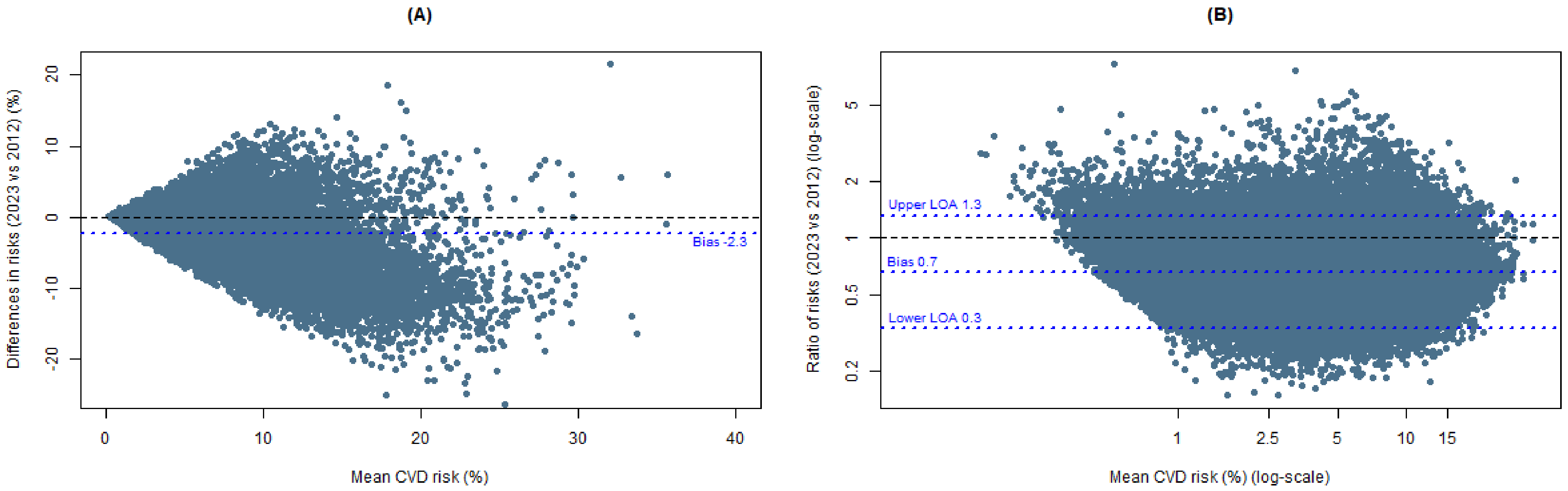
